# Stroke Risk Reduction in Migraine Patients Using Propranolol: Evidence from Two Large-Scale Real-World Data Analyses

**DOI:** 10.1101/2024.06.11.24308801

**Authors:** Eugene Jeong, Mulubrhan F. Mogos, You Chen

## Abstract

**Background:** Propranolol, a non-selective beta-blocker, is commonly used for migraine prevention, but its impact on stroke risk among migraine patients remains controversial. Using two large electronic health records-based datasets, we examined stroke risk differences between migraine patients with- and without- documented use of propranolol

**Methods:** This retrospective case-control study utilized EHR data from the Vanderbilt University Medical Center (VUMC) and the All of Us Research Program. Migraine patients were first identified based on the International Classification of Headache Disorders, 3rd edition (ICHD-3) criteria using diagnosis codes. Among these patients, cases were defined as those with a primary diagnosis of stroke following the first diagnosis of migraine, while controls had no stroke after their first migraine diagnosis. Logistic regression models, adjusted for potential factors associated with stroke risk, assessed the association between propranolol use and stroke risk, stratified by sex and migraine subtype. A Cox proportional hazards regression model was used to estimate the hazard ratio (HR) for stroke risk at 1, 2, 5, and 10 years from baseline.

**Results:** In the VUMC database, 378 cases and 15,209 controls were identified, while the All of Us database included 267 cases and 6,579 controls. Propranolol significantly reduced stroke risk in female migraine patients (VUMC: OR=0.52, p=0.006; All of Us: OR=0.39, *p*=0.007), but not in males. The effect was more pronounced for ischemic stroke and in females with migraines without aura (MO) (VUMC: OR=0.60, p=0.014; All of Us: OR=0.28, p=0.006). The Cox model showed lower stroke rates in propranolol-treated female migraine patients at 1, 2, 5, and 10 years (VUMC: HR=0.06-0.55, p=0.0018-0.085; All of Us: HR=0.23, p=0.045 at 10 years).

**Conclusions:** Propranolol is associated with a significant reduction in stroke risk, particularly ischemic stroke, among female migraine without aura patients. These findings suggest that propranolol may benefit stroke prevention in high-risk populations.

## Introduction

Migraine, a neurological disorder characterized by recurrent headaches, affects a substantial portion of the global population, significantly impacting quality of life and productivity^1^. Growing evidence suggests that migraine can increase the risk of stroke, an acute and often catastrophic cerebrovascular event that causes significant physical disability and mortality worldwide^2–4^. Epidemiological studies have shown that migraine is associated with an increased risk of both ischemic^5–10^ and hemorrhagic strokes^7,11–13^. For instance, the relative risk of ischemic stroke is doubled in individuals with migraine with aura compared to those without migraine^5,7,8,10^, with the risk being greater for those experiencing active migraine attacks in the past 12 months and a higher frequency of attacks^6,9,14^. Additionally, there is an increased risk of hemorrhagic stroke among migraine patients, especially in women under 45^12^. Given the association of migraine with the risk of stroke, exploring preventive strategies for stroke in high-risk groups such as migraine patients is crucial. At present, there are no pharmacologic interventions specifically recommended for stroke prevention in migraine patients, underscoring the need for research to determine whether currently used migraine prophylactic medications can mitigate the risk of stroke.

Propranolol, a non–selective beta–blocker, is widely used in the prevention of migraines^15^. Its efficacy in migraine prevention is well–documented^15,16^, but its potential role in the association of migraine with the risk of stroke remains controversial ^17–22^. Some studies suggest that propranolol may paradoxically increase the risk of stroke due to its effects on cerebral blood flow and blood pressure regulation^20–22^, while others find no significant association or even a protective effect^17–19^. The controversy is further complicated by the heterogeneity of study designs, patient populations, and outcome measures^21,22^. This inconsistency presents a significant challenge in drawing definitive conclusions and highlights the need for more robust, controlled studies.

The purpose of this study is to evaluate the potential role of propranolol in reducing the risk of stroke in migraine patients through a comprehensive analysis of clinical data from two large electronic health records (EHR).

## Methods

### Data Sources

This retrospective case-control study was conducted using two anonymized EHR databases: the Synthetic Derivative (SD) maintained by Vanderbilt University Medical Center (VUMC)^23^ and the All of Us Research Program managed by the National Institutes of Health (NIH)^24^. The SD database comprises de-identified longitudinal research data of over 3 million individuals spanning more than 15 years. The All of Us Research Program has EHR data for over 230,000 diverse participants across the United States as of May 2024. Notably, the two EHR databases could not be merged due to potential patient record overlap. The study received approval from the Institutional Review Board (IRB) at VUMC, under approval identifier #221125.

### Exposure: Migraine Patient Identification

Patients diagnosed with migraine were selected based on the International Classification of Headache Disorders, 3rd edition (ICHD–3)^25^, which serves as the gold standard for migraine diagnosis, particularly in research contexts (**Figure 1**). Migraines were classified into migraine with aura (MA), migraine without aura (MO), and unclassified migraine using the International Classification of Diseases, Ninth Revision (ICD-9), and Tenth Revision (ICD-10) codes. For patients with MA, identification was based on ICD9 codes 346.0, 346.01, 346.02, 346.03, 346.5x, and 346.6x, along with ICD10 codes G43.1x, with at least one encounter required. Patients with MO were identified using ICD9 codes 346.1, 346.11, 346.12, 346.13, and 346.7x, as well as ICD10 codes G43.0x, with at least four encounters required. Patients whose migraines did not fit neatly into MA or MO categories were considered as unclassified migraine patients and were identified using ICD9 codes 346.2x, 346.8x, and 346.9x, and ICD10 codes G43.8x and G43.9x, with at least two encounters required. For exclusion criteria, patients with coexistent brain disorders or conditions (ICD9: 340–344.99, 347–349.99; ICD10: G35.x, G36.x, G47., G80.x, G81.x, G82.x, G93.x) and brain tumors (ICD9: 191.x, 192.x, 239.6, 239.7; ICD10: C71, C72, D49.6, D49.7) were excluded. Since the ICHD-3 notes that children’s migraines are often bilateral and shorter in duration compared to adults, patients under 18 at migraine diagnosis were excluded. The complete list of ICD codes for migraine conditions is provided in **Table S1**.

**Figure 1.**
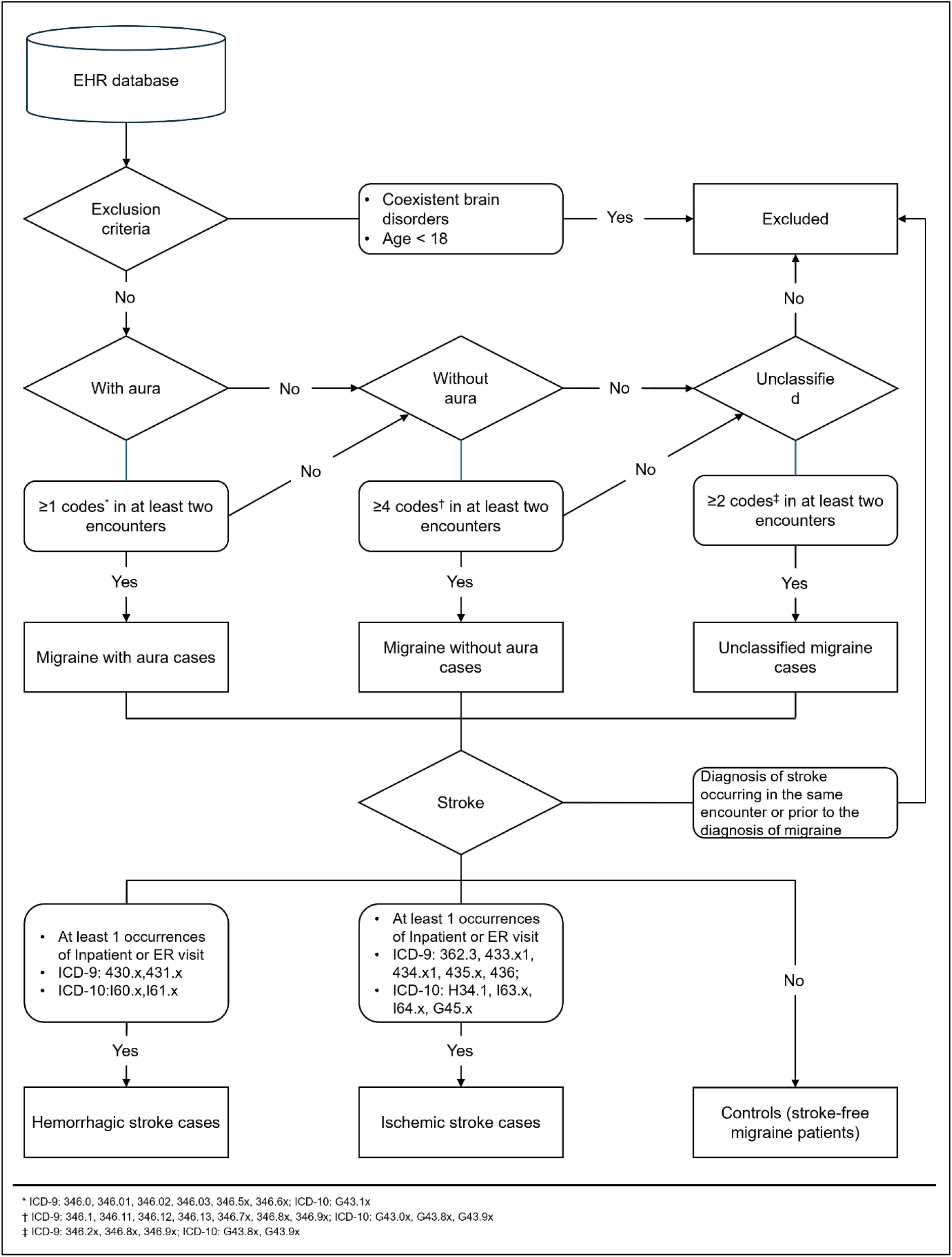
Flowchart illustrating the process of defining cases and controls among migraine patients.

### Outcomes: Stroke Patient Identification

Stroke cases were defined as having a stroke event requiring inpatient admission with a primary diagnosis of major stroke types, including ischemic stroke (acute ischemic stroke [AIS] and transient ischemic attack [TIA]) or hemorrhagic stroke (intracerebral hemorrhage [ICH] and subarachnoid hemorrhage [SAH]) (**Figure 1**). AIS was identified using ICD-9 codes 362.3, 433.x1, 434.x1, 436, and ICD-10 codes H34.1, I63.x, and I64.x. TIA was identified using ICD-9 code 435.x and ICD-10 code G45.x. ICH was identified with ICD-9 code 431.x and ICD-10 code I61.x. SAH was identified with ICD-9 code 430.x and ICD-10 code I60.x. The complete list of ICD codes for stroke conditions is provided in **Table S2**.

Cases were defined as individuals with a history of stroke after migraine diagnosis; controls had no history of stroke after the diagnosis of migraine. Patients were excluded if they had stroke codes prior to migraine diagnosis or were missing age/sex information. We considered three types of strokes throughout the study: hemorrhagic, ischemic, and overall strokes. Notably, the overall stroke category may exceed the sum of hemorrhagic and ischemic strokes because some patients were diagnosed with both hemorrhagic and ischemic stroke during the same encounter. In such cases, we excluded these patients when investigating the risk of hemorrhagic stroke or ischemic stroke individually but included them when assessing the risk of overall strokes.

### Covariates

All potential factors associated with stroke risk were identified and defined based on Leppert et al^2^. These factors were categorized into demographic, comorbidity, hormonal, and treatment factors. Demographic factors included variables such as age, sex, and race. Comorbidity factors encompassed conditions such as hypertension (including gestational hypertension), diabetes (including gestational diabetes), hyperlipidemia, sleep apnea, peripheral artery disease, atrial fibrillation, coronary artery disease, alcohol abuse, substance abuse, tobacco use, obesity, congestive heart failure, malignancy, HIV, hepatitis, thrombophilia (including history of deep vein thrombosis and pulmonary embolism), autoimmune disease, vasculitis, sickle cell disease, heart valve disease, and renal failure. Hormonal factors, including the use of oral contraceptives and pregnancy, were considered separately for women. Treatment factors involved first-line migraine medications such as valproate, topiramate, metoprolol, timolol, and methysergide.

The Healthcare Cost and Utilization Project (HCUP) Clinical Classification Software was used to identify comorbidity and hormonal factors using ICD-9 and ICD-10 codes^26^. For factors not covered by HCUP, such as sleep apnea, atrial fibrillation, tobacco use, pregnancy, and vasculitis, validated ICD-9 and ICD-10 codes identified through literature review were used. Pharmacy claims were used to identify the use of migraine treatments and oral contraceptives in women based on the anatomical therapeutic chemical (ATC) classification. Additionally, pharmacy claims supplemented the diagnosis of diabetes, dyslipidemia, migraines, malignancy, and HIV. The comorbidity status was assessed by determining if migraine patients had any comorbid conditions before the diagnosis of migraines. Treatment history was evaluated by checking if migraine patients received treatments between the diagnosis of migraines and the diagnosis of strokes for cases and after the diagnosis of migraines for controls. To verify pregnancy status for cases, we first identified all women who had ICD-9 or ICD-10 codes indicating a pregnancy outcome, such as delivery, spontaneous abortion, or elective abortion. We then determined the start and end dates of the pregnancy to check if the stroke date occurred during the pregnancy or postpartum period. To ensure that no pregnancy-related strokes were missed due to the subscriber moving out of state before delivery, we also searched for any antenatal care codes within 9 months prior to the stroke date. For controls, we checked whether individuals were pregnant or in the postpartum period at or after the diagnosis of the migraine date. A detailed list of covariates including ICD-9 and ICD-10 codes are provided in **Table S3**.

## Statistical Analysis

We analyzed the risk of each stroke type in overall migraine patients. Additionally, we examined the risk of each stroke type in specific migraine subtypes to assess whether propranolol’s effect varies among them.

We compared baseline characteristics between cases and controls using Fisher’s exact test, unpaired t-test, or Wilcoxon rank-sum test.

The association between propranolol and stroke risk was evaluated using univariate and multivariate logistic regression analyses, adjusting for covariates and stratifying by sex. Statistical significance was defined as p < 0.05 and an odds ratio (OR) < 1.

To assess the association between stroke incidence rates and propranolol use, cumulative event rates were calculated using the Kaplan-Meier method. A covariate-adjusted Cox proportional hazards regression model was fitted to estimate the hazard ratios (HR), 2-sided 95% confidence intervals (CI), and 2-sided Wald p-values (with p < 0.05 considered significant) at 1, 2, 5, and 10 years. All statistical analyses were conducted using R software.

## Results

The VUMC SD database comprised 273,491,202 diagnosis records and 868,741,933 drug prescription records for 3,183,571 patients, spanning from January 1989 to May 2023. In contrast, the All of Us EHR database contained 86,012,976 diagnosis records and 79,531,217 drug prescription records for 239,715 patients, covering the period from January 1981 to July 2022. Among these, there were 15,587 eligible migraine patients in the VUMC database and 6,846 eligible migraine patients in the All of Us EHR database (**Figure 2**).

**Figure 2.**
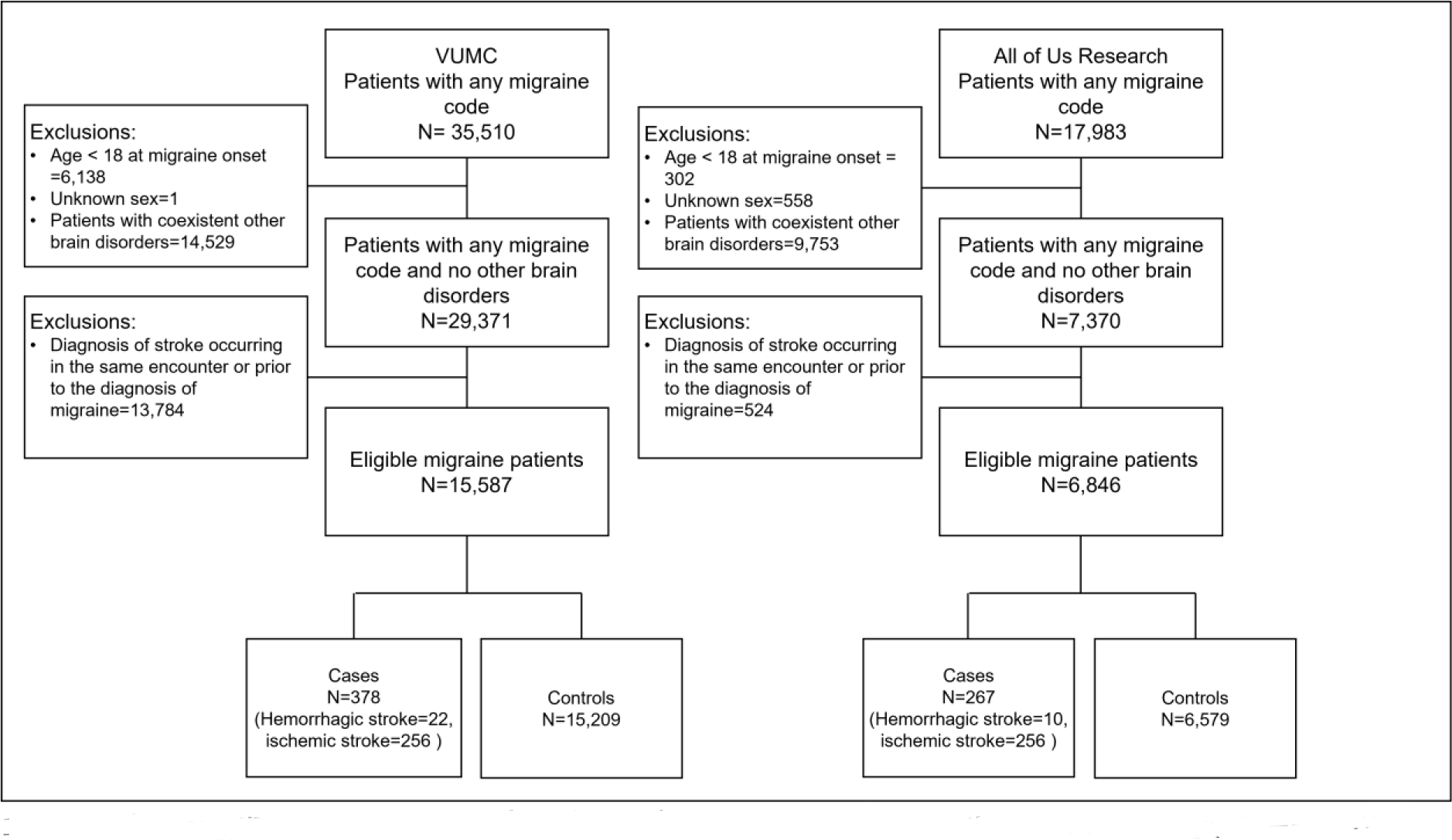
Eligibility and inclusion flowchart from the VUMC and All of Us Research EHR databases.

In the VUMC EHR database, there were 378 cases (82% female; 67.7% ischemic stroke) and 15,209 controls (**Figure 2**). Among men, the most common comorbidities in cases were hyperlipidemia (44%), malignancy (19%), and hypertension (18%), while in women, they were hyperlipidemia (28%), hypertension (18%), and malignancy (16%) (**Table 1**). Women with stroke were as likely to be pregnant (cases: 9.4%, controls: 9.4%) but less likely to use oral contraception compared to controls (cases: 20%, controls: 28%). The All of Us Research EHR database contained 267 cases (80.9% women; 95.9% ischemic strokes) and 6,579 controls (**Figure 2**). In both men and women, the most common comorbidities among cases were hyperlipidemia (35% in men, 36% in women), hypertension (20% in men, 30% in women), and diabetes (16% in men, 19% in women) (**Table 1**). Women who had a stroke were less likely to be pregnant (cases: 0.5%, controls: 2.9%) and less likely to use oral contraception compared to controls (cases: 11%, controls: 18%).

**Table 1.** Summary of descriptive statistics summarizing patient demographic and clinical characteristics by sex for cases and controls in the VUMC and All of Us EHR databases.

| Variables | VUMC |  |  |  |  |  | All of Us |  |  |  |  |  |
| --- | --- | --- | --- | --- | --- | --- | --- | --- | --- | --- | --- | --- |
|  | Male |  |  | Female |  |  | Male |  |  | Female |  |  |
|  | Cases<br>N = 68 | Controls<br>N = 2,306 | p-value<br>† | Cases<br>N = 310 | Controls<br>N = 12,903 | p-value<br>† | Cases<br>N = 51 | Controls<br>N = 797 | p-value<br>† | Cases<br>N = 216 | Controls<br>N = 5,782 | p-value<br>† |
| **Demographic factors** |  |  |  |  |  |  |  |  |  |  |  |  |
| Age | 49.8 (14.9) | 42.7 (14.7) | <0.001 | 48.7 (15.2) | 40.0 (13.9) | <0.001 | 51.7 (13.0) | 46.1 (15.1) | 0.005 | 50.4 (14.3) | 42.3 (14.1) | <0.001 |
| Race |  |  |  |  |  |  |  |  |  |  |  |  |
| White | 59 (87%) | 1,907 (83%) | 0.297 | 262 (85%) | 10,563 (82%) | 0.212 | 36 (71%) | 528 (66%) | 0.970 | 141 (65%) | 3,579 (62%) | 0.724 |
| American Indian or Alaska Native | 1 (1.5%) | 3 (0.1%) |  | 1 (0.3%) | 17 (0.1%) |  | 0 (0%) | 0 (0%) |  | 0 (0%) | 0 (0%) |  |
| Asian | 0 (0%) | 36 (1.6%) |  | 2 (0.6%) | 132 (1.0%) |  | 1 (2.0%) | 18 (2.3%) |  | 1 (0.5%) | 93 (1.6%) |  |
| Black or African American | 4 (5.9%) | 152 (6.6%) |  | 30 (9.7%) | 1,217 (9.4%) |  | 6 (12%) | 96 (12%) |  | 24 (11%) | 730 (13%) |  |
| Middle Eastern or North African | 0 (0%) | 4 (0.2%) |  | 1 (0.3%) | 16 (0.1%) |  | 0 (0%) | 7 (0.9%) |  | 1 (0.5%) | 28 (0.5%) |  |
| Native Hawaiian or Other Pacific Islander | 0 (0%) | 1 (<0.1%) |  | 0 (0%) | 4 (<0.1%) |  | 0 (0%) | 0 (0%) |  | 0 (0%) | 6 (0.1%) |  |
| Unknown | 4 (5.9%) | 203 (8.8%) |  | 14 (4.5%) | 954 (7.4%) |  | 8 (16%) | 148 (19%) |  | 49 (23%) | 1,346 (23%) |  |
| **Comorbidity factors** |  |  |  |  |  |  |  |  |  |  |  |  |
| Hypertension | 12 (18%) | 244 (11%) | 0.073 | 57 (18%) | 1,131 (8.8%) | <0.001 | 10 (20%) | 205 (26%) | 0.407 | 65 (30%) | 1,171 (20%) | <0.001 |
| Complicated hypertension | 3 (4.4%) | 50 (2.2%) | 0.192 | 14 (4.5%) | 203 (1.6%) | <0.001 | 0 (0%) | 28 (3.5%) | 0.405 | 8 (3.7%) | 150 (2.6%) | 0.280 |
| Diabetes | 11 (16%) | 200 (8.7%) | 0.047 | 38 (12%) | 1,170 (9.1%) | 0.058 | 8 (16%) | 113 (14%) | 0.684 | 42 (19%) | 751 (13%) | 0.010 |
| Complicated diabetes | 4 (5.9%) | 58 (2.5%) | 0.099 | 16 (5.2%) | 219 (1.7%) | <0.001 | 2 (3.9%) | 29 (3.6%) | 0.709 | 16 (7.4%) | 174 (3.0%) | 0.001 |
| Hyperlipidemia | 30 (44%) | 459 (20%) | <0.001 | 88 (28%) | 1,765 (14%) | <0.001 | 18 (35%) | 270 (34%) | 0.879 | 77 (36%) | 1,309 (23%) | <0.001 |
| Sleep apnea | 1 (1.5%) | 75 (3.3%) | 0.724 | 13 (4.2%) | 443 (3.4%) | 0.431 | 2 (3.9%) | 40 (5.0%) | >0.999 | 12 (5.6%) | 272 (4.7%) | 0.514 |
| Peripheral artery disease | 2 (2.9%) | 45 (2.0%) | 0.392 | 6 (1.9%) | 118 (0.9%) | 0.072 | 2 (3.9%) | 11 (1.4%) | 0.181 | 6 (2.8%) | 121 (2.1%) | 0.465 |
| Atrial fibrillation | 2 (2.9%) | 34 (1.5%) | 0.276 | 2 (0.6%) | 75 (0.6%) | 0.703 | 2 (3.9%) | 23 (2.9%) | 0.658 | 8 (3.7%) | 58 (1.0%) | 0.002 |
| Coronary artery disease | 11 (16%) | 82 (3.6%) | <0.001 | 11 (3.5%) | 218 (1.7%) | 0.024 | 5 (9.8%) | 64 (8.0%) | 0.598 | 16 (7.4%) | 200 (3.5%) | 0.008 |
| Alcohol use disorder | 0 (0%) | 9 (0.4%) | >0.999 | 2 (0.6%) | 29 (0.2%) | 0.164 | 0 (0%) | 25 (3.1%) | 0.393 | 2 (0.9%) | 37 (0.6%) | 0.651 |
| Substance use disorder | 7 (10%) | 117 (5.1%) | 0.086 | 15 (4.8%) | 423 (3.3%) | 0.145 | 6 (12%) | 119 (15%) | 0.685 | 21 (9.7%) | 413 (7.1%) | 0.179 |
| Tobacco use | 3 (4.4%) | 113 (4.9%) | >0.999 | 11 (3.5%) | 451 (3.5%) | 0.876 | 8 (16%) | 135 (17%) | >0.999 | 17 (7.9%) | 629 (11%) | 0.180 |
| Obesity | 1 (1.5%) | 74 (3.2%) | 0.723 | 22 (7.1%) | 974 (7.5%) | 0.913 | 4 (7.8%) | 72 (9.0%) | >0.999 | 18 (8.3%) | 920 (16%) | 0.002 |
| Congestive heart failure | 1 (1.5%) | 23 (1.0%) | 0.504 | 3 (1.0%) | 93 (0.7%) | 0.494 | 1 (2.0%) | 13 (1.6%) | 0.583 | 4 (1.9%) | 67 (1.2%) | 0.325 |
| Malignancy | 13 (19%) | 237 (10%) | 0.027 | 50 (16%) | 1,516 (12%) | 0.026 | 3 (5.9%) | 82 (10%) | 0.468 | 25 (12%) | 684 (12%) | >0.999 |
| HIV | 1 (1.5%) | 26 (1.1%) | 0.546 | 0 (0%) | 24 (0.2%) | >0.999 | 1 (2.0%) | 10 (1.3%) | 0.497 | 1 (0.5%) | 27 (0.5%) | >0.999 |
| Hepatitis | 1 (1.5%) | 22 (1.0%) | 0.489 | 1 (0.3%) | 45 (0.3%) | >0.999 | 1 (2.0%) | 26 (3.3%) | >0.999 | 6 (2.8%) | 83 (1.4%) | 0.138 |
| Thrombophilia | 4 (5.9%) | 79 (3.4%) | 0.298 | 21 (6.8%) | 447 (3.5%) | 0.004 | 3 (5.9%) | 46 (5.8%) | >0.999 | 18 (8.3%) | 324 (5.6%) | 0.099 |
| Autoimmune diseases | 1 (1.5%) | 30 (1.3%) | 0.596 | 21 (6.8%) | 358 (2.8%) | <0.001 | 2 (3.9%) | 17 (2.1%) | 0.318 | 10 (4.6%) | 299 (5.2%) | 0.875 |
| Vasculitis | 0 (0%) | 5 (0.2%) | >0.999 | 1 (0.3%) | 34 (0.3%) | 0.565 | 0 (0%) | 4 (0.5%) | >0.999 | 2 (0.9%) | 33 (0.6%) | 0.361 |
| Sickle cell disease | 0 (0%) | 9 (0.4%) | >0.999 | 1 (0.3%) | 48 (0.4%) | >0.999 | 0 (0%) | 2 (0.3%) | >0.999 | 2 (0.9%) | 44 (0.8%) | 0.682 |
| Heart valve disorder | 7 (10%) | 67 (2.9%) | 0.005 | 17 (5.5%) | 356 (2.8%) | 0.008 | 3 (5.9%) | 47 (5.9%) | >0.999 | 28 (13%) | 368 (6.4%) | <0.001 |
| Renal failure | 4 (5.9%) | 62 (2.7%) | 0.118 | 7 (2.3%) | 167 (1.3%) | 0.131 | 0 (0%) | 48 (6.0%) | 0.108 | 8 (3.7%) | 137 (2.4%) | 0.251 |
| **Hormonal factors** |  |  |  |  |  |  |  |  |  |  |  |  |
| Pregnancy |  |  |  | 29 (9.4%) | 1,209 (9.4%) | >0.999 |  |  |  | 1 (0.5%) | 167 (2.9%) | 0.032 |
| Oral contraceptives |  |  |  | 62 (20%) | 3,658 (28%) | 0.001 |  |  |  | 23 (11%) | 1,012 (18%) | 0.008 |
| **Migraine treatment factors** |  |  |  |  |  |  |  |  |  |  |  |  |
| Valproate | 7 (10%) | 225 (9.8%) | 0.836 | 18 (5.8%) | 1,096 (8.5%) | 0.098 | 1 (2.0%) | 48 (6.0%) | 0.354 | 8 (3.7%) | 192 (3.3%) | 0.699 |
| Topiramate | 8 (12%) | 420 (18%) | 0.202 | 60 (19%) | 3,314 (26%) | 0.012 | 7 (14%) | 107 (13%) | >0.999 | 35 (16%) | 1,196 (21%) | 0.122 |
| Metoprolol | 9 (13%) | 279 (12%) | 0.708 | 30 (9.7%) | 1,255 (9.7%) | >0.999 | 5 (9.8%) | 134 (17%) | 0.242 | 36 (17%) | 726 (13%) | 0.077 |
| Timolol | 1 (1.5%) | 22 (1.0%) | 0.489 | 1 (0.3%) | 82 (0.6%) | >0.999 | 0 (0%) | 17 (2.1%) | 0.617 | 4 (1.9%) | 78 (1.3%) | 0.541 |
| Methysergide | 0 (0%) | 1 (<0.1%) | >0.999 | 0 (0%) | 2 (<0.1%) | >0.999 | 0 (0%) | 1 (0.1%) | >0.999 | 0 (0%) | 1 (<0.1%) | >0.999 |
| Propranolol | 6 (8.8%) | 261 (11%) | 0.696 | 19 (6.1%) | 1,604 (12%) | <0.001 | 2 (3.9%) | 78 (9.8%) | 0.218 | 9 (4.2%) | 618 (11%) | <0.001 |

Mean (SD); n (%)
†Unpaired t-test or Welch rank-sum test; Fisher's exact test

Potential factors with no cases were inestimable and excluded from both univariate and multivariate models. In the VUMC EHR database, alcohol use disorder, vasculitis, sickle cell disease, and methysergide were excluded from the male group, while methysergide was excluded from the female group. propranolol was not significantly associated with the risk of overall stroke in men according to the covariate-adjusted multivariate model (OR=0.93, p=0.88), but it was significantly associated with a reduced risk of overall stroke in women (OR=0.52, p=0.006) (**Table 2** **and Table S4**). In the All of Us Research EHR database, complicated hypertension, alcohol use disorder, vasculitis, sickle cell disease, renal failure, timolol, and methysergide were excluded from the male group, while methysergide was excluded from the female group. Propranolol was not significantly associated with the risk of overall stroke in men according to the covariate-adjusted multivariate model (OR=0.52, p=0.39), whereas it was significantly associated with a reduced risk of overall stroke in women (OR=0.39, p=0.007) (**Table 2** **and Table S4**).

**Table 2.** Multivariate logistic regression analysis in a case-control study of migraine patients by sex within the VUMC and All of Us EHR databases.

| Variables | VUMC |  |  |  | All of Us |  |  |  |
| --- | --- | --- | --- | --- | --- | --- | --- | --- |
|  | Male |  | Female |  | Male |  | Female |  |
|  | AOR<br>(95% CI) | p-value | AOR<br>(95% CI) | p-value | AOR<br>(95% CI) | p-value | AOR<br>(95% CI) | p-value |
| **Demographic factors** |  |  |  |  |  |  |  |  |
| Age | 1.01<br>(1.0-1.03) | 0.15 | 1.03<br>(1.03-1.04) | <0.001 | 1.03<br>(1.01-1.06) | 0.007 | 1.03<br>(1.02-1.04) | <0.001 |
| Race |  |  |  |  |  |  |  |  |
| White | Referent | Referent | Referent | Referent | Referent | Referent | Referent | Referent |
| American Indian or Alaska Native | 23.5<br>(0.82-347) | 0.024 | 2.22<br>(0.12-11.3) | 0.44 | Nim* | Nim* | Nim* | Nim* |
| Asian | Nim* | Nim* | 0.70<br>(0.11-2.24) | 0.62 | 0.81<br>(0.04-4.76) | 0.85 | 0.28<br>(0.02-1.29) | 0.21 |
| Black or African American | 1.08<br>(0.31-2.80) | 0.89 | 1.21<br>(0.79-1.77) | 0.36 | 1.00<br>(0.34-2.52) | >0.99 | 1.01<br>(0.61-1.59) | 0.97 |
| Middle Eastern or North African | Nim* | Nim* | 2.38<br>(0.13-13.0) | 0.42 | Nim* | Nim* | 1.21<br>(0.07-5.98) | 0.86 |
| Native Hawaiian or Other Pacific Islander | Nim* | Nim* | Nim* | Nim* | Nim* | Nim* | Nim* | Nim* |
| Unknown | 0.81<br>(0.24-2.04) | 0.69 | 0.66<br>(0.36-1.09) | 0.13 | 0.93<br>(0.38-2.03) | 0.85 | 1.08<br>(0.76-1.53) | 0.65 |
| **Comorbidity factors** |  |  |  |  |  |  |  |  |
| Hypertension | 0.91<br>(0.36-2.07) | 0.84 | 1.57<br>(1.07-2.27) | 0.018 | 0.55<br>(0.22-1.24) | 0.17 | 1.19<br>(0.82-1.69) | 0.35 |
| Complicated Hypertension | 0.47<br>(0.07-2.51) | 0.41 | 1.54<br>(0.75-3.02) | 0.22 | Nim* | Nim* | 0.98<br>(0.39-2.19) | 0.96 |
| Diabetes | 0.85<br>(0.31-2.03) | 0.74 | 0.73<br>(0.45-1.13) | 0.18 | 1.23<br>(0.40-3.27) | 0.70 | 1.15<br>(0.71-1.78) | 0.56 |
| Complicated Diabetes | 1.68<br>(0.37-6.83) | 0.48 | 2.57<br>(1.27-5.09) | 0.007 | 1.14<br>(0.14-6.29) | 0.89 | 2.36<br>(1.17-4.66) | 0.014 |
| Hyperlipidemia | 2.42<br>(1.28-4.50) | 0.006 | 1.40<br>(1.03-1.90) | 0.031 | 0.76<br>(0.36-1.57) | 0.47 | 1.10<br>(0.78-1.55) | 0.58 |
| Sleep Apnea | 0.35<br>(0.02-1.75) | 0.32 | 0.92<br>(0.48-1.60) | 0.78 | 0.87<br>(0.13-3.40) | 0.86 | 1.00<br>(0.51-1.80) | 0.99 |
| Peripheral Artery Disease | 0.47<br>(0.07-1.97) | 0.37 | 0.93<br>(0.34-2.10) | 0.87 | 2.88<br>(0.35-17.1) | 0.27 | 0.63<br>(0.23-1.44) | 0.32 |
| Atrial Fibrillation | 0.52<br>(0.06-2.69) | 0.49 | 0.45<br>(0.07-1.57) | 0.29 | 2.35<br>(0.33-10.5) | 0.31 | 2.65<br>(1.03-6.08) | 0.030 |
| Coronary Artery Disease | 5.77<br>(1.59-20.1) | 0.006 | 0.78<br>(0.31-1.80) | 0.58 | 2.18<br>(0.38-11.5) | 0.36 | 0.86<br>(0.38-1.89) | 0.71 |
| Alcohol Use Disorder | Nim* | Nim* | 2.87<br>(0.42-11.3) | 0.19 | Nim* | Nim* | 1.41<br>(0.22-5.16) | 0.65 |
| Substance Use Disorder | 0.51<br>(0.11-2.12) | 0.37 | 1.07<br>(0.46-2.43) | 0.86 | 0.55<br>(0.11-2.21) | 0.43 | 1.46<br>(0.70-2.99) | 0.30 |
| Tobacco Use | 0.61<br>(0.12-2.14) | 0.49 | 0.74<br>(0.33-1.50) | 0.44 | 1.47<br>(0.55-3.51) | 0.41 | 0.62<br>(0.33-1.09) | 0.12 |
| Obesity | 0.25<br>(0.01-1.34) | 0.20 | 0.71<br>(0.43-1.12) | 0.16 | 1.08<br>(0.30-3.00) | 0.89 | 0.38<br>(0.22-0.63) | <0.001 |
| Congestive Heart Failure | 0.26<br>(0.01-1.99) | 0.28 | 0.44<br>(0.10-1.34) | 0.20 | 3.75<br>(0.16-37.0) | 0.30 | 0.71<br>(0.19-2.06) | 0.56 |
| Malignancy | 1.36<br>(0.63-2.70) | 0.41 | 0.93<br>(0.66-1.30) | 0.69 | 0.53<br>(0.12-1.65) | 0.33 | 0.64<br>(0.39-0.99) | 0.052 |
| HIV | 1.22<br>(0.06-7.32) | 0.86 | 0.00<br>(0.00-28.3) | 0.97 | 2.22<br>(0.11-15.3) | 0.48 | 1.40<br>(0.08-6.97) | 0.74 |
| Hepatitis | 1.46<br>(0.07-9.52) | 0.74 | 0.62<br>(0.03-3.19) | 0.65 | 0.87<br>(0.04-5.24) | 0.90 | 1.63<br>(0.60-3.73) | 0.29 |
| Thrombophilia | 0.80<br>(0.19-2.61) | 0.74 | 1.56<br>(0.93-2.50) | 0.075 | 0.94<br>(0.18-3.41) | 0.93 | 1.40<br>(0.81-2.31) | 0.20 |
| Autoimmune Diseases | 0.96<br>(0.05-5.54) | 0.97 | 2.01<br>(1.20-3.20) | 0.005 | 3.22<br>(0.44-14.6) | 0.17 | 0.67<br>(0.32-1.26) | 0.25 |
| Vasculitis | Nim* | Nim* | 0.71<br>(0.04-3.63) | 0.75 | Nim* | Nim* | 1.49<br>(0.23-5.50) | 0.61 |
| Sickle Cell Disease | Nim* | Nim* | 0.81<br>(0.04-4.02) | 0.83 | Nim* | Nim* | 1.94<br>(0.30-6.81) | 0.38 |
| Heart Valve Disorder | 2.72<br>(0.94-6.88) | 0.047 | 1.38<br>(0.77-2.33) | 0.25 | 0.59<br>(0.11-2.21) | 0.48 | 1.80<br>(1.13-2.77) | 0.010 |
| Renal Failure | 1.24<br>(0.27-4.47) | 0.77 | 0.64<br>(0.25-1.44) | 0.32 | Nim* | Nim* | 0.83<br>(0.34-1.82) | 0.67 |
| **Hormonal factors** |  |  |  |  |  |  |  |  |
| Pregnancy |  |  | 1.50<br>(0.96-2.27) | 0.061 |  |  | 0.28<br>(0.02-1.30) | 0.21 |
| Oral Contraceptives |  |  | 0.67<br>(0.49-0.91) | 0.011 |  |  | 0.87<br>(0.53-1.36) | 0.55 |
| **Migraine treatment factors** |  |  |  |  |  |  |  |  |
| Valproate | 1.27<br>(0.50-2.77) | 0.58 | 0.83<br>(0.49-1.32) | 0.46 | 0.35<br>(0.02-1.80) | 0.32 | 1.56<br>(0.68-3.10) | 0.25 |
| Topiramate | 0.65<br>(0.28-1.34) | 0.28 | 0.81<br>(0.60-1.07) | 0.15 | 1.37<br>(0.53-3.10) | 0.48 | 0.92<br>(0.62-1.33) | 0.68 |
| Metoprolol | 0.73<br>(0.31-1.53) | 0.44 | 0.72<br>(0.47-1.05) | 0.10 | 0.51<br>(0.17-1.27) | 0.19 | 1.05<br>(0.70-1.53) | 0.80 |
| Timolol | 1.03<br>(0.05-5.94) | 0.98 | 0.25<br>(0.01-1.19) | 0.18 | Nim* | Nim* | 0.89<br>(0.27-2.24) | 0.83 |
| Methysergide | Nim* | Nim* | Nim* | Nim* | Nim* | Nim* | Nim* | Nim* |
| Propranolol | 0.93<br>(0.35-2.08) | 0.88 | 0.52<br>(0.31-0.80) | 0.006 | 0.52<br>(0.08-1.85) | 0.39 | 0.39<br>(0.18-0.73) | 0.007 |

Nim, not in the model; OR, odds ratio; CI, confidence intervals; AOR, adjusted odds ratio
\* Due to low numbers of factors within the sex category (case=0), some factor estimates were inestimable and thus not included in the model

In the stratified analysis by stroke type, a significant reduction in the odds of ischemic stroke associated with propranolol was observed only in females across both EHR databases based on the covariate-adjusted multivariate model (**Table S5– S8**). In the stratified analysis by migraine type, a significant reduction in the odds of overall and ischemic stroke associated with propranolol was observed only in females with MO based on the covariate-adjusted multivariate model in both EHR databases (**Table S9–S14**).

Given the observed reduction in the odds of stroke associated with propranolol only in females across both EHR databases, we further assessed the HR in female groups only. Potential factors with no cases were excluded from the covariate-adjusted Cox proportional hazards regression model. According to the VUMC database, the cumulative incidence of stroke was numerically lower in propranolol-treated female migraine patients at each time point: 0.25% vs 1.31% at 1 year, 0.37% vs 1.55% at 2 years, 0.43% vs 1.87% at 5 years, and 0.92% vs 2.15% at 10 years. The covariate-adjusted HRs were 0.06 (95% CI:0.016-0.23; p=0.0018), 0.22 (95% CI:0.083-0.57; p=0.018), 0.55 (95% CI:0.23-1.3; p=0.085), and 0.44 (95% CI:0.24-0.83; p=0.0064) at 1, 2, 5, and 10 years, respectively (**Figure 3**), indicating a lower risk of stroke compared to those not taking propranolol.

**Figure 3.**
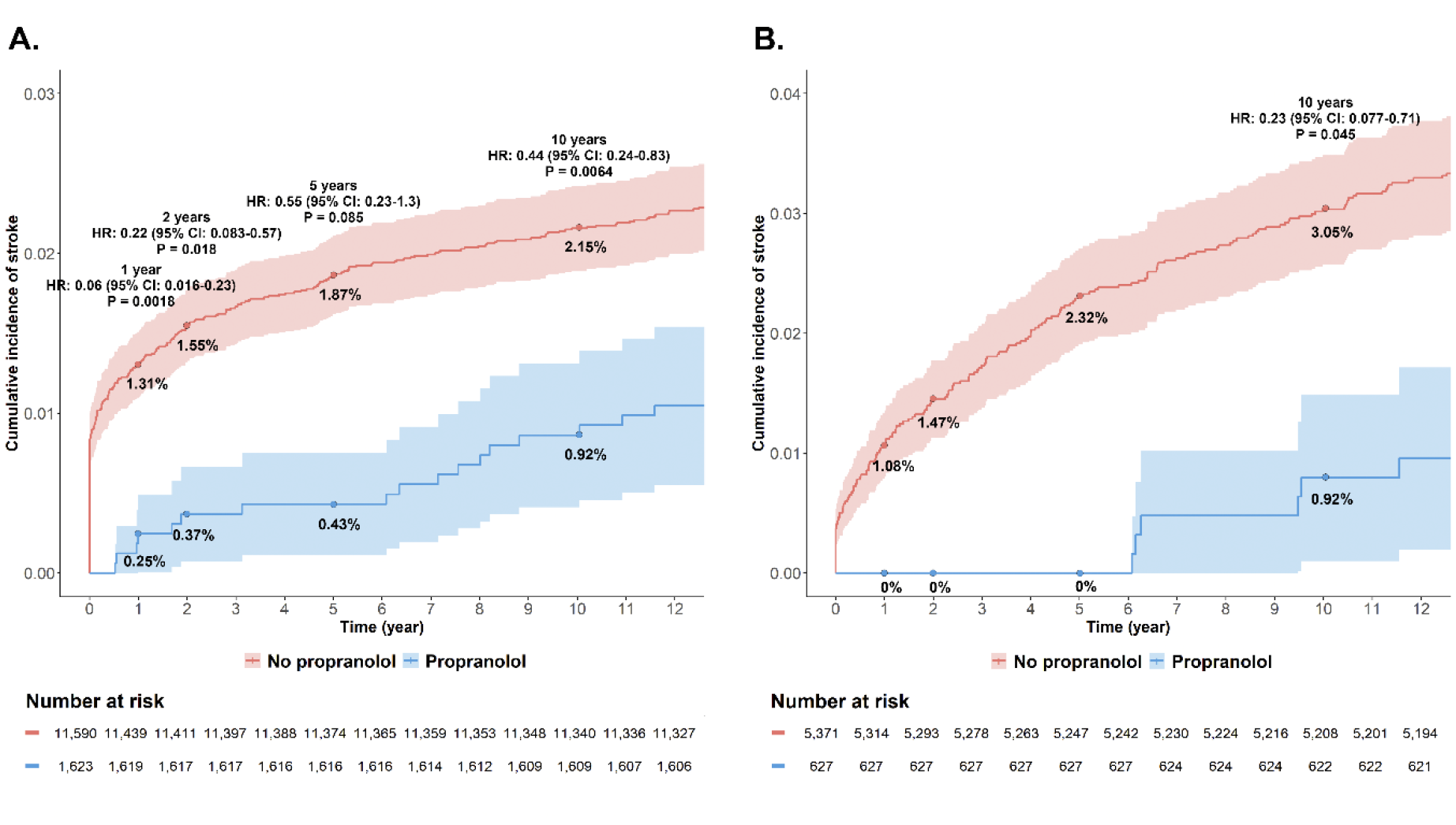
Cumulative Incidence of Stroke Events in Female Migraine Patients. (A) Kaplan-Meier curves with adjusted hazard ratios (HR) and 95% confidence intervals (CI) at 1, 2, 5, and 10 years from the VUMC database. (B) Kaplan-Meier curves with adjusted HRs and 95% CIs at 1, 2, 5, and 10 years from the All of Us database.

In the All of Us database, the cumulative incidence of stroke was also numerically lower in propranolol-treated female migraine patients at each time point: 0% vs 1.08% at 1 year, 0% vs 1.47% at 2 years, 0% vs 2.32% at 5 years, and 0.92% vs 3.05% at 10 years. Since there were no stroke events in the propranolol-treated group at 1, 2, and 5 years, the covariate-adjusted HRs could not be calculated for those time points, but the covariate-adjusted HR at 10 years was 0.23 (95% CI:0.077-0.72; p=0.045) (**Figure 3**).

## Discussion

This study provides significant insights into the potential role of propranolol in reducing the risk of stroke among migraine patients, with a particular focus on sex-specific effects. The consistency of the findings across two large and diverse EHR databases adds credibility to the results. The VUMC database is more region-specific with a potentially homogeneous population, while the All of Us database includes a broader, more heterogeneous population representative of various regions of the United States. Despite differences in patient demographics and clinical characteristics between the VUMC and All of Us cohorts, the association between propranolol and reduced stroke risk in females remained significant, particularly for ischemic stroke. The significant reduction in stroke risk could be attributed to propranolol’s effects on blood pressure and heart rate variability (HRV). Propranolol is effective in managing high blood pressure, a major risk factor for ischemic stroke. By lowering blood pressure, propranolol reduces the strain on blood vessels, thereby preventing vascular events such as strokes^27,28^. The drug also influences HRV by enhancing parasympathetic activity and stabilizing heart rate, which can lead to improved cardiovascular health and a reduced risk of stroke^29^.

The study highlights a noteworthy sex difference in the association between propranolol and the risk of ischemic stroke. In both databases, propranolol use was associated with a significant reduction in the risk of overall and ischemic strokes in female migraine patients, but not in males. The enhanced protective effect against stroke in female patients could be attributed to sex-specific variations in drug metabolism and hormonal influences. Previous research indicates that women exhibit higher peak plasma levels and area under the plasma concentration-time curve (AUC) for propranolol compared to men, due to enhanced absorption, reduced volume of distribution, and slower clearance through Cytochrome P450 2D6 (CYP2D6)^30–32^. These factors lead to a more significant reduction in heart rate and systolic blood pressure during exercise for women^30–32^. These pharmacokinetic differences can enhance the drug’s efficacy in reducing cardiovascular risks for females. Hormonal influences, particularly estrogen, may also amplify propranolol’s protective effects. Estrogen has been found to modulate vascular tone and improve endothelial function, which, in combination with propranolol’s beta-blocking properties, could synergistically lower stroke risk^33^. Moreover, previous studies have shown that the fluctuations in CGRP-mediated trigeminovascular responses over the menstrual cycle indicated a potential connection between sex hormones and the trigeminovascular system^2^, highlighting the significance of taking into account gender-specific aspects in migraine research and treatment^30^.

The analysis indicates that the protective effect of propranolol is more significant in female patients with MO. This subtype-specific effect suggests that the pathophysiological mechanisms linking migraines and stroke risk may vary by migraine subtype, potentially influencing the efficacy of prophylactic treatments like propranolol. Primarily associated with dysregulation of the central nervous system, MO involves less cortical spreading depression compared to MA^34^. This difference in pathophysiology might make MO more responsive to propranolol which stabilizes neural activity and reduces systemic cardiovascular risks. Moreover, propranolol’s effectiveness in reducing stroke risk in MO patients could also be attributed to its ability to modulate sympathetic nervous system activity, which is more prominently dysregulated in MO. The lack of aura in MO patients means there is a lower baseline risk of stroke due to fewer episodes of cortical spreading depression (CSD), which is a known precipitant of ischemic stroke in MA patients^34^. This makes the cardiovascular protective effects of propranolol more apparent in the MO group. Further research is necessary to elucidate these mechanisms and to understand why propranolol appears more beneficial for certain migraine subtypes.

Several limitations must be acknowledged. First, the factors were identified retrospectively based on an administrative dataset, so we cannot account for factors present in controls or cases whose diagnoses were not coded. Additionally, the retrospective design limits the ability to infer causality. Second, relying on ICD codes for migraine and stroke diagnoses may introduce misclassification bias. Finally, although the study included a large number of patients, the number of stroke events, particularly hemorrhagic strokes, was relatively low, potentially limiting the power to detect significant associations in this subgroup.

Future research should focus on prospective studies to confirm these findings and to explore the underlying mechanisms of propranolol’s protective effects against stroke. Personalized medicine approaches that tailor stroke prevention strategies based on individual risk profiles and migraine characteristics could further optimize patient outcomes.

## Data Availability

The VUMC SD dataset is not publicly available. The All of Us Research EHR database is owned by a third party, the All of Us Research Program.

